# Essential Emergency and Critical Care – a consensus among global clinical experts

**DOI:** 10.1101/2021.03.18.21253191

**Authors:** Carl Otto Schell, Karima Khalid, Alexandra Wharton-Smith, Jacquie Narotso Oliwa, Hendry Robert Sawe, Nobhojit Roy, Alex Sanga, John C. Marshall, Jamie Rylance, Claudia Hanson, Raphael Kazidule Kayambankadzanja, Lee A. Wallis, Maria Jirwe, Tim Baker, the EECC Collaborators

## Abstract

**Background:** Globally, critical illness results in millions of deaths every year. Although many of these deaths are potentially preventable, the basic, life-saving care of critically ill patients are often overlooked in health systems. Essential Emergency and Critical Care (EECC) has been devised as the care that should be provided to all critically ill patients in all hospitals in the world. EECC includes the effective care of low cost and low complexity for the identification and timely treatment of critically ill patients across all medical specialities. This study aimed to specify the content of EECC and additionally, given the surge of critical illness in the ongoing pandemic, the essential diagnosis-specific care for critically ill patients with COVID-19.

**Methods:** A Delphi process was conducted to seek consensus (>90% agreement) in a diverse panel of global clinical experts. The panel was asked to iteratively rate proposed treatments and actions based on previous guidelines and the WHO/ICRC’s Basic Emergency Care. The output from the Delphi was adapted iteratively with specialist reviewers into a coherent and feasible EECC package of clinical processes plus a list of hospital resource requirements.

**Results:** The 269 experts in the Delphi panel had clinical experience in different acute medical specialties from 59 countries and from all resource settings. The agreed EECC package contains 40 clinical processes and 67 hospital readiness requirements. The essential diagnosis-specific care of critically ill COVID-19 patients has an additional 7 clinical processes and 9 hospital readiness requirements.

**Conclusion:** The study has specified the content of the essential emergency and critical care that should be provided to all critically ill patients. Implementation of EECC could be an effective strategy to reduce preventable deaths worldwide. As critically ill patients have high mortality rates, especially where trained staff or resources are limited, even small improvements would have a large impact on survival. EECC has a vital role in the effective scale-up of oxygen and other care for critically ill patients in the COVID-19 pandemic. Policy makers should prioritise EECC, increase its coverage in hospitals, and include EECC as a component of universal health coverage.

## Introduction

Critical illness, when defined as a state of ill health with vital organ dysfunction and a high risk of imminent death, is common in hospitals throughout the world [1-6]. It is the most severe form of acute illness due to any underlying condition and results in millions of deaths globally every year [1, 5]. The COVID-19 pandemic has led to increased morbidity and mortality with a surge in critical illness worldwide [7-9].

Many of the deaths due to critical illness are potentially preventable [10-12]. In critical illness, the patient’s airway, breathing, or circulation may become compromised, and early identification of the problem and timely care can be lifesaving. Unfortunately, this care is frequently a neglected part of healthcare. The basic, life-saving clinical processes may be overlooked in specialised care [12] and in settings of both high [13-15] and low resources [16-18]. In hospitals all over the world, guidelines, equipment, and routines focusing on the care of critically ill patients, are often missing for adult[19] and paediatric patients[11], in emergency units [20], in wards [21] and in intensive care units [22]. Improving the way healthcare manages critical illness could save many lives [11, 23, 24].

To improve outcomes for critically ill patients by means that are feasible to deliver in all hospital wards and settings, the Essential Emergency and Critical Care (EECC) concept was devised [25]. EECC is defined as the care that should be provided to all critically ill patients of all ages in all hospitals in the world. It is distinguished by three principles. First, priority to those with the most urgent clinical need, including both early identification and timely care. Second, provision of the life-saving treatments that support and stabilise failing vital organ functions. And third, a focus on effective care of low cost and low complexity.

The clinical processes that comprise the essential care of critically ill patients, and the resources required for those processes have not previously been specified. As critically ill patients can be suffering from any underlying condition, EECC is conceptualised to be integrated into all acute clinical specialties. We therefore sought consensus among a diverse group of global clinical experts with the aim of specifying the content of EECC. An additional aim, given the ongoing pandemic, was consensus around the essential diagnosis-specific care for critically ill patients with COVID-19.

## Methods

The study used three phases. (**Figure 1**) Firstly, a consensus was sought about the treatments and actions (T&A) in EECC using a modified Delphi technique [26]. Secondly, the output from the Delphi was adjusted into a coherent, user-friendly, and feasible package of clinical processes. And thirdly, a list of requirements for hospitals to be ready to provide the care was developed.

**Figure 1.**
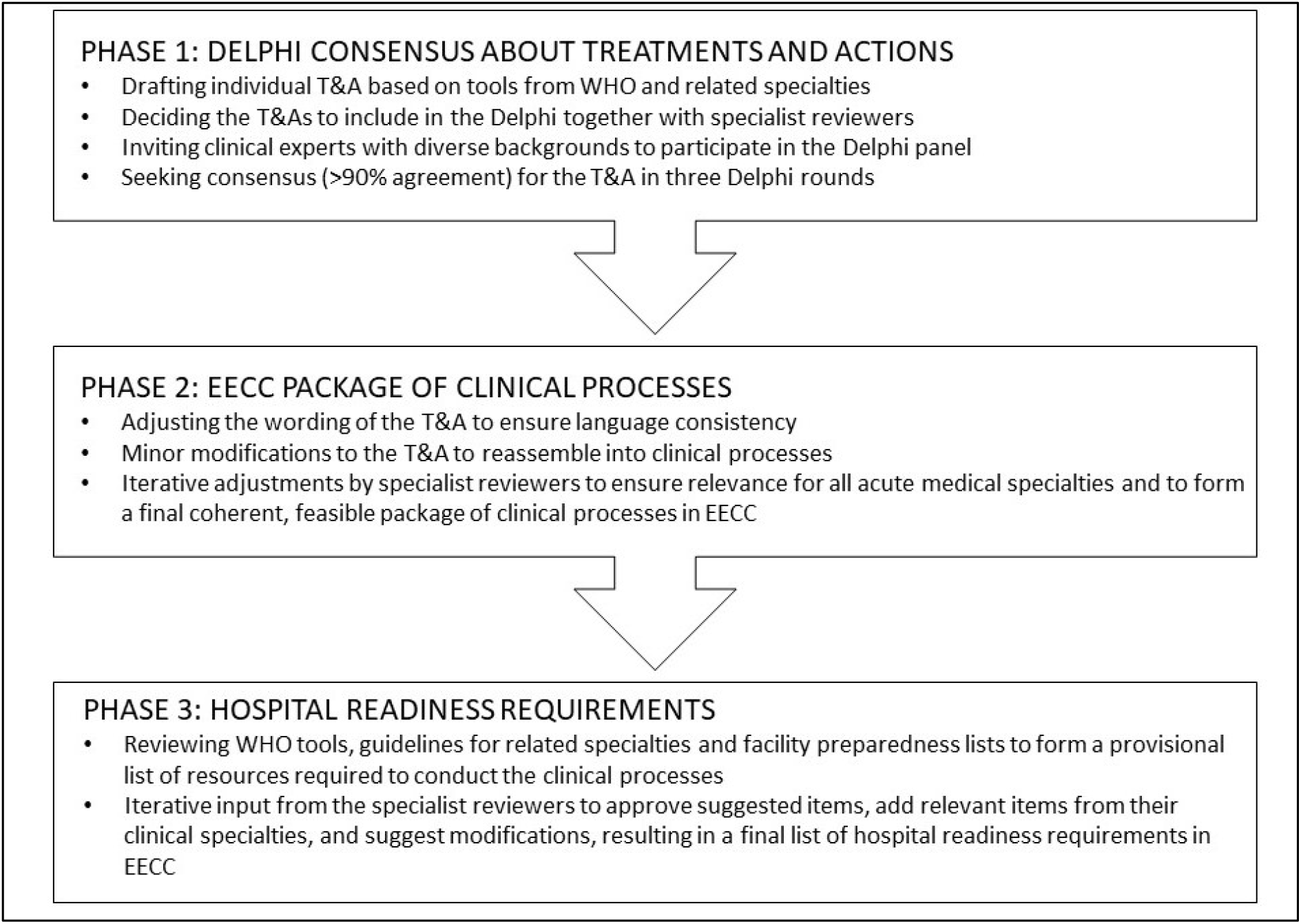
Summary of the process. *EECC* Essential Emergency and critical care *T&A* Treatment and actions *WHO* World Health Organisation

### Phase One

An online, three-round modified Delphi process was conducted in November and December 2020. The Delphi method uses anonymous responses from an expert panel to iteratively posed questions and controlled feedback to reach consensus on the topic of interest [26]. A Delphi process was chosen for this study as EECC is new, its content has not been previously specified and a large group of diverse experts was required.

To be part of the panel, experts needed to have clinical experience of caring for critically ill patients. To ensure the involvement of a diverse range of experts, it was decided that at least 50% of the invitations to participate in the panel should be sent to experts with substantial experience of working in low- and middle-income countries, and there should be a balance between clinical experience (work in general wards, emergency units, intensive care units); specialty (paediatrics, obstetrics, medicine, surgery, intensive care, anaesthesia and emergency care); profession (doctors, nurses, other health professionals); location, and gender. A list of potential participants was made from a mapping of stakeholders, the literature across all acute medical specialties, the researchers’ networks and additional purposive and snowball sampling for under-represented groups. Additionally, a link to a screening survey was sent to global professional networks, specialist societies and on social media to identify further potential participants. A total of 895 experts were invited to participate, and those who accepted provided written informed consent.

EECC consists of clinical processes of care. To enable rating by the Delphi panel, clinical processes were disassembled into individual treatments & actions (T&A). The T&A concern the identification of critical illness; care of critical illness, and the diagnosis-specific care of critically ill COVID-19 patients. To be included, all T&A were required to meet two a-priori defined criteria: *effectiveness*\* and *feasibility*\*. Additionally, *universality*\* was required for the identification and care of critical illness and *relevance*\* was required for the diagnosis-specific care of critically ill patients with COVID-19. A draft list of potential T&A was developed based on clinical guidelines and tools from related specialties [27-38] and aligned with the WHO/ICRC’s Basic Emergency Care [39]. The draft list was revised by specialist reviewers – a group of senior clinicians, researchers, and policy makers, with expertise in paediatrics, medicine, emergency medicine, anaesthesia and intensive care, critical care nursing, obstetrics and gynaecology, and surgery.

Three Delphi rounds were deemed sufficient to address the aim while avoiding attrition and poor response rates. A four-point Likert scale (strongly disagree, disagree, agree, strongly agree) with a ‘do-not know’ option was used for the panel to rate their opinion about the inclusion of each T&A in EECC [40-42]. Consensus was achieved when more than 90% of respondents selected “agree” or “strongly agree”, excluding “don’t know” responses. The experts were able to provide free-text comments, which were analysed to identify appropriate, relevant changes to the wording of T&A for clarity of understanding, and to identify newly proposed T&A. After the first round, newly proposed T&A that fulfilled the EECC criteria for potential inclusion were revised after input from the specialist reviewers and included for assessment by the panel. T&A that did not reach consensus in the previous round were presented for re-assessment in rounds two and three, together with a visual representation of the spread of previous responses.

As the Delphi panel was diverse, it was considered that there may be different opinions about the inclusion of T&As between experts with particular a-priori defined characteristics. These subgroups of experts were those with work experience in a low-income country or not; those who are doctors or not; those with clinical experience in emergency care and those without; and those with clinical experience in intensive care and those without. The levels of agreement in each subgroup were assessed and presented for all the T&As that reached consensus.

### Phase Two

After the Delphi, slight adjustments were made to the wording of the T&A that had reached consensus to ensure language consistency. The T&A were reassembled back into clinical processes to increase overall coherence and feasibility of the EECC package, with the goal of user-friendliness for health system implementation and quality improvement work. The adjustments were done in an iterative process with the same specialist reviewers as in Phase One to ensure relevance for all acute medical specialties. The final package of clinical processes was organised into those relevant for identification, for care, and general processes.

### Phase Three

A provisional list of hospital readiness requirements for the provision of the clinical processes were developed using existing WHO tools, guidelines for related specialties, facility preparedness lists [29, 32, 34, 35, 37-39, 43, 44] and the experience and knowledge of the study team. The specialist reviewers provided iterative input into the provisional list, approving suggested items, adding relevant items from their clinical specialties, and suggesting modifications. Based on previous work and following consultation with health economists and procurement experts, the final list of requirements was agreed and arranged into eight categories: equipment, consumables, drugs, human resources, training, routines, guidelines, and infrastructure.

## Results

### Phase One

Of the 895 invited experts, 269 participated in the first round of the Delphi when the majority of the decisions were made (30% response rate). In Round Two, 228 experts participated (85% of those in Round One) and Round Three included 194 experts (85% of those in Round Two). The panel comprised experts from diverse resource settings, clinical settings, specialties, and professions **(Table 1)**. The panel included experts from 59 countries (**Figure 2)** and 38% were female.

**Table 1.** The characteristics of the expert panel in the Delphi (first round)

|  | Number of experts (N=269) | Proportion of experts (%) |
| --- | --- | --- |
| **Resource setting\*** |  |  |
| High-income country | 139 | 52 |
| Middle-income country | 115 | 43 |
| Low-income country | 177 | 66 |
| Do not know | 2 | 1 |
| **Clinical setting\*** |  |  |
| General ward | 153 | 57 |
| Emergency unit | 179 | 67 |
| High dependency unit | 153 | 57 |
| Intensive care unit | 232 | 86 |
| Operating theatre | 102 | 38 |
| Other | 15 | 6 |
| **Specialty\*** |  |  |
| Emergency Care | 93 | 35 |
| Intensive Care | 190 | 71 |
| Anaesthesia | 59 | 22 |
| Medicine | 39 | 15 |
| Surgery | 20 | 7 |
| Paediatrics | 47 | 17 |
| Obstetrics/Gynaecology | 13 | 5 |
| Other | 25 | 9 |
| **Profession\*** |  |  |
| Doctor | 212 | 79 |
| Nurse | 40 | 15 |
| Midwife | 6 | 2 |
| Clinical Officer | 9 | 3 |
| Other | 17 | 6 |
| **Gender** |  |  |
| Female | 102 | 38 |
| Male | 165 | 62 |

\*As the experts were asked to select all that apply, the sum of the percentages may exceed 100

**Figure 2.**
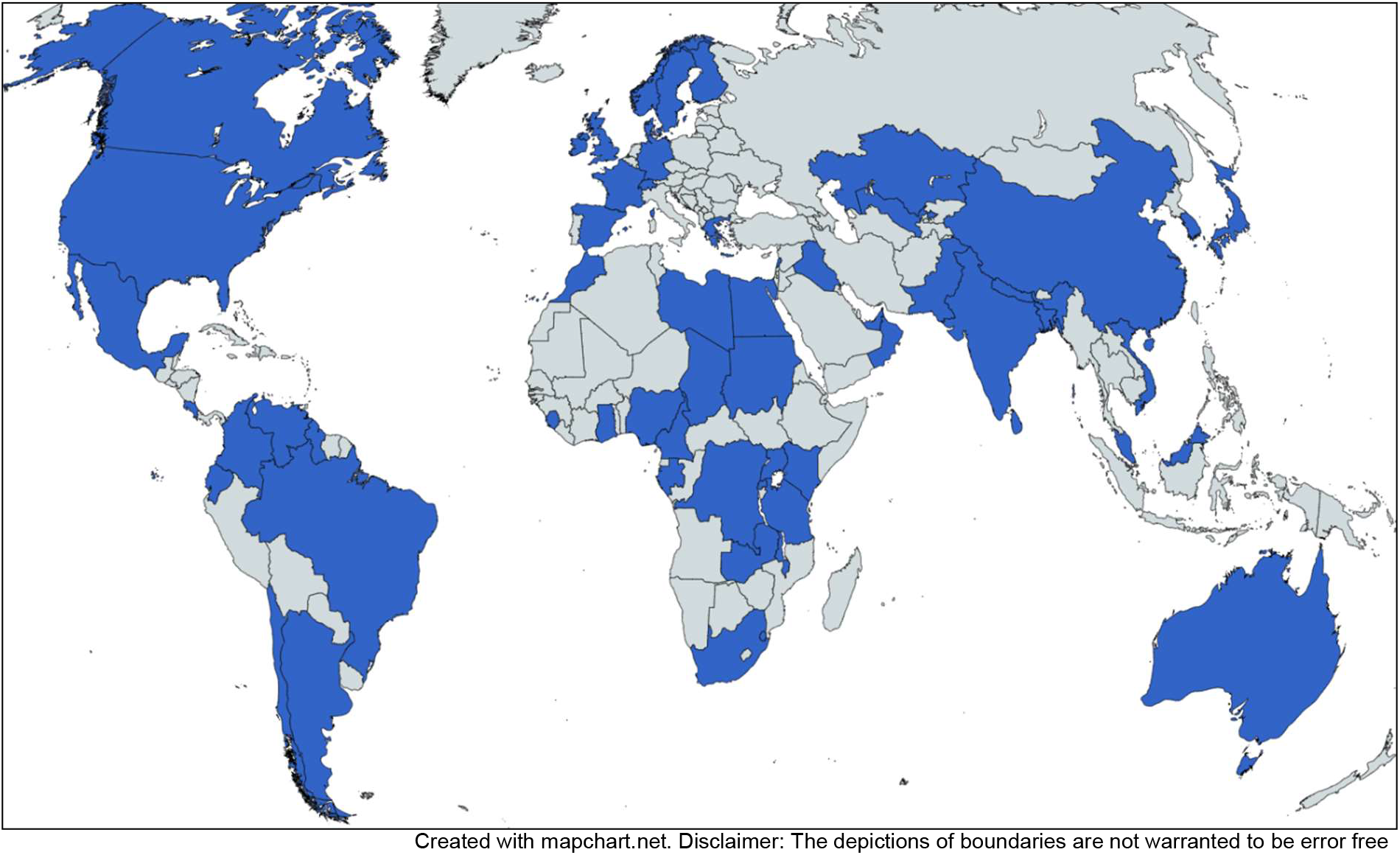
Expert panel locations. The locations of the expert panel in the Delphi Round One

Of the 57 T&A for EECC in Round One, consensus was reached for 49. In Round Two, 29 newly proposed T&A were added to the eight remaining from Round One, of which two had been re-worded for clarity. Out of these 37, consensus was reached for 17. The remaining 20, of which another two had been re-worded for clarity, were included in Round Three. Consensus was reached for nine of the final 20 T&A. In total, consensus was reached for 75 out of 86 proposed T&A, including 54 of the original 57. (**Supplementary table 1**)

Of the seven T&A for the essential diagnosis-specific care of critically ill COVID-19 in Round One, all reached consensus for inclusion. In Round Two, two newly proposed T&A were added. Neither of these reached consensus in Round Two or Round Three.

Analyses of participant sub-groups did not reveal substantial divergence from the overall results. For the T&A that reached 90% agreement in the panel, agreement was not below 80% in any subgroup. (**Supplementary tables 2-4)**

### Phase Two and Three

After the Delphi, the T&A that had reached consensus were reassembled into a final user-friendly and feasible package of EECC containing 40 clinical processes – 30 identification and care processes and 10 general processes. (**Panel 1**) All T&A for the care of critical illness were included, with some rewording and reordering. Eleven T&A for the identification of critical illness were not included, so that the package could be feasible for triage in all hospitals, and were added as an addendum (outside the remit of EECC), in order to underscore their importance in settings where staff have sufficient time and expertise.

The list of hospital readiness requirements for EECC contained 67 items, (fourteen for identification and 53 for essential care). **(Panel 2)**

The essential diagnosis-specific care of critically ill COVID-19 patients consisted of an additional seven clinical processes and nine hospital readiness requirements. (**Panel 3**)

**PANEL 1.**
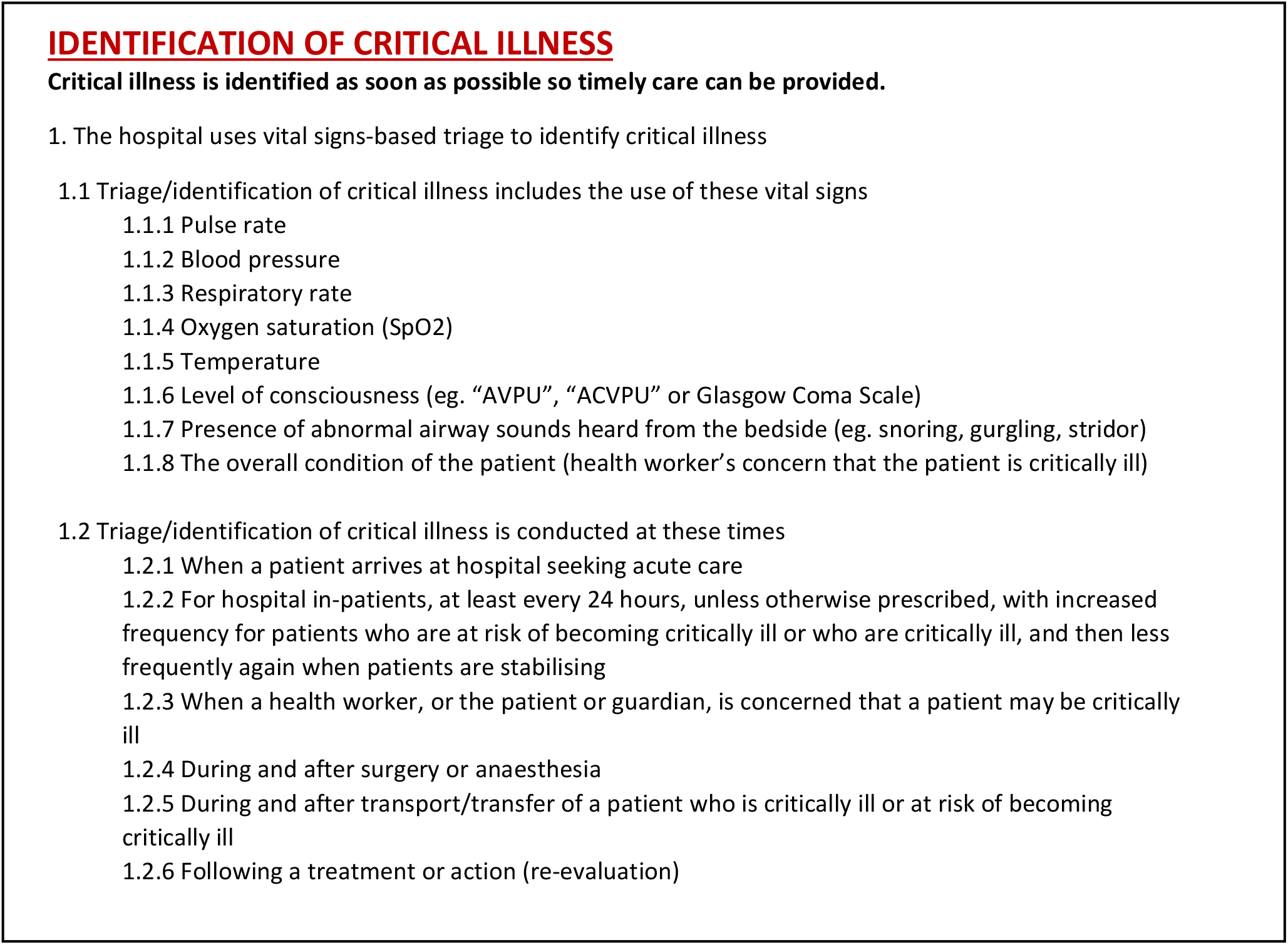
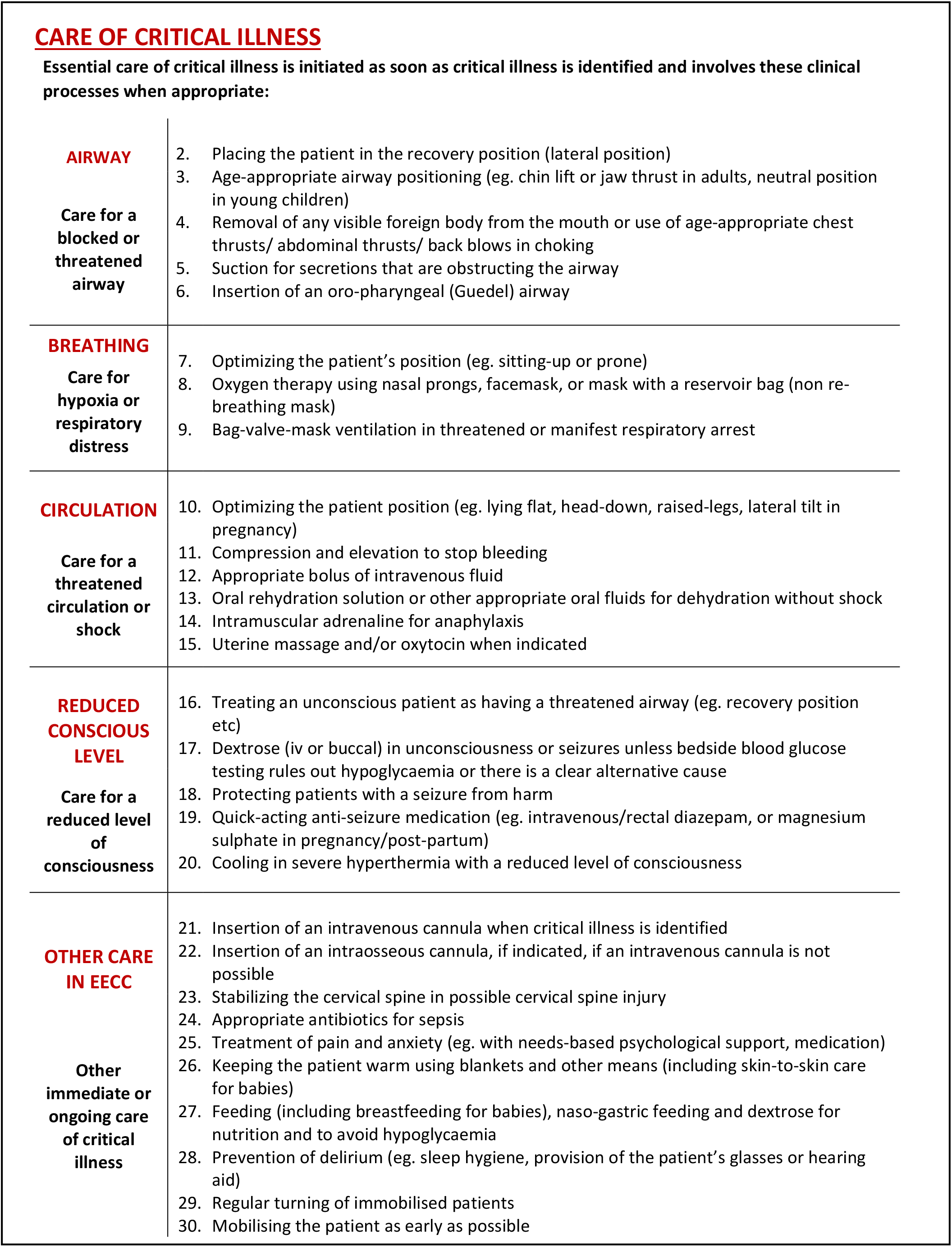
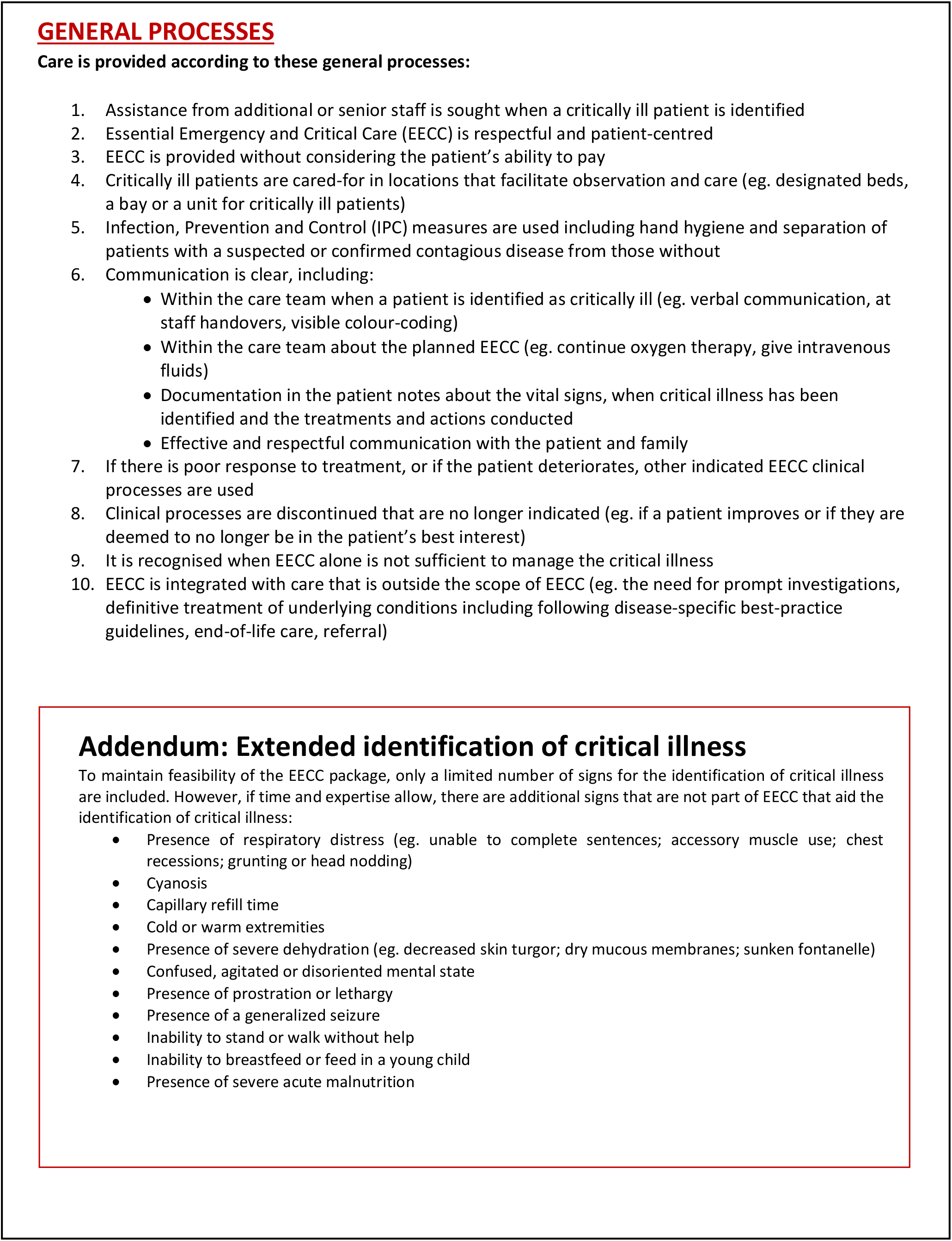
The clinical processes of Essential Emergency and Critical Care.

**PANEL 2.**
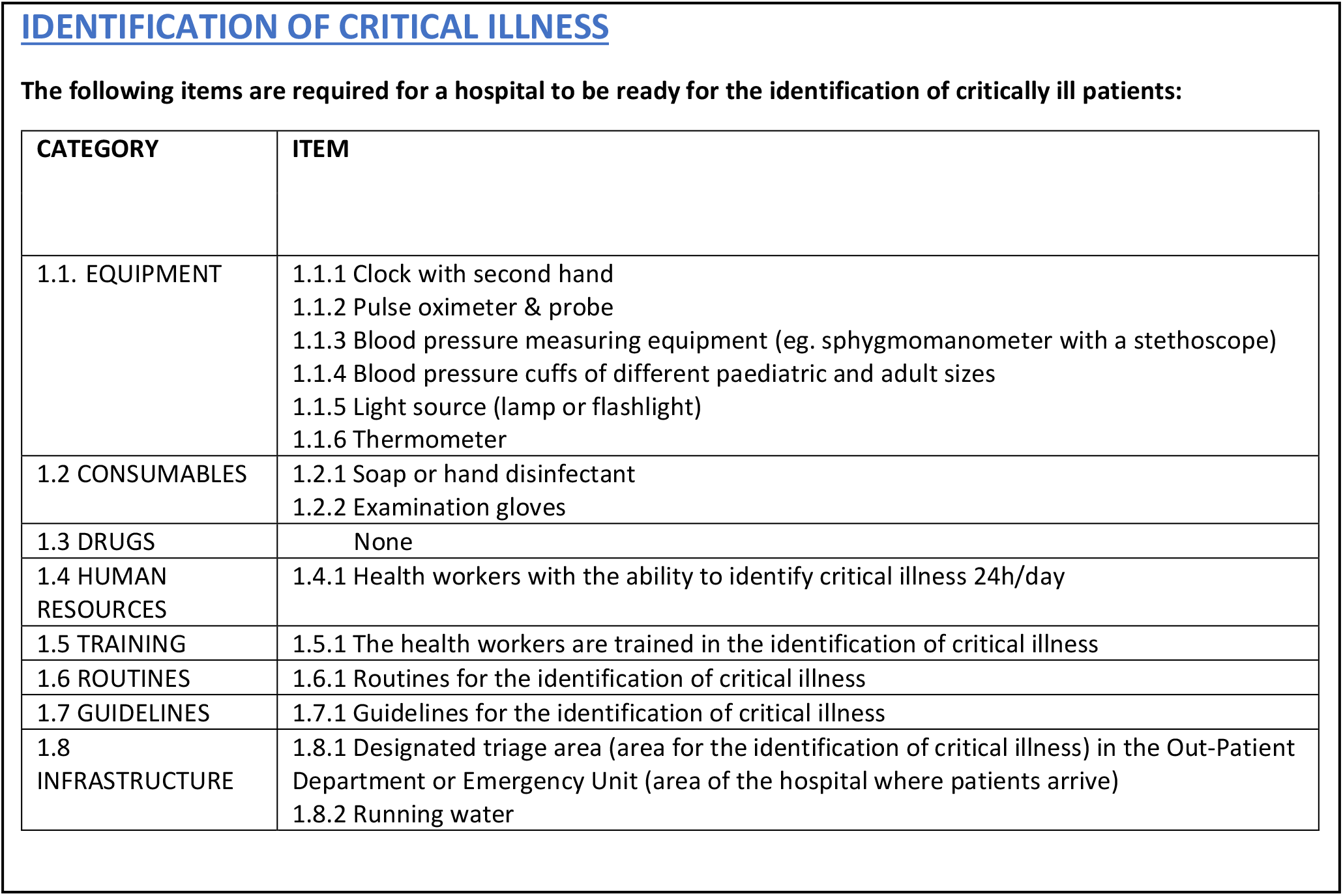
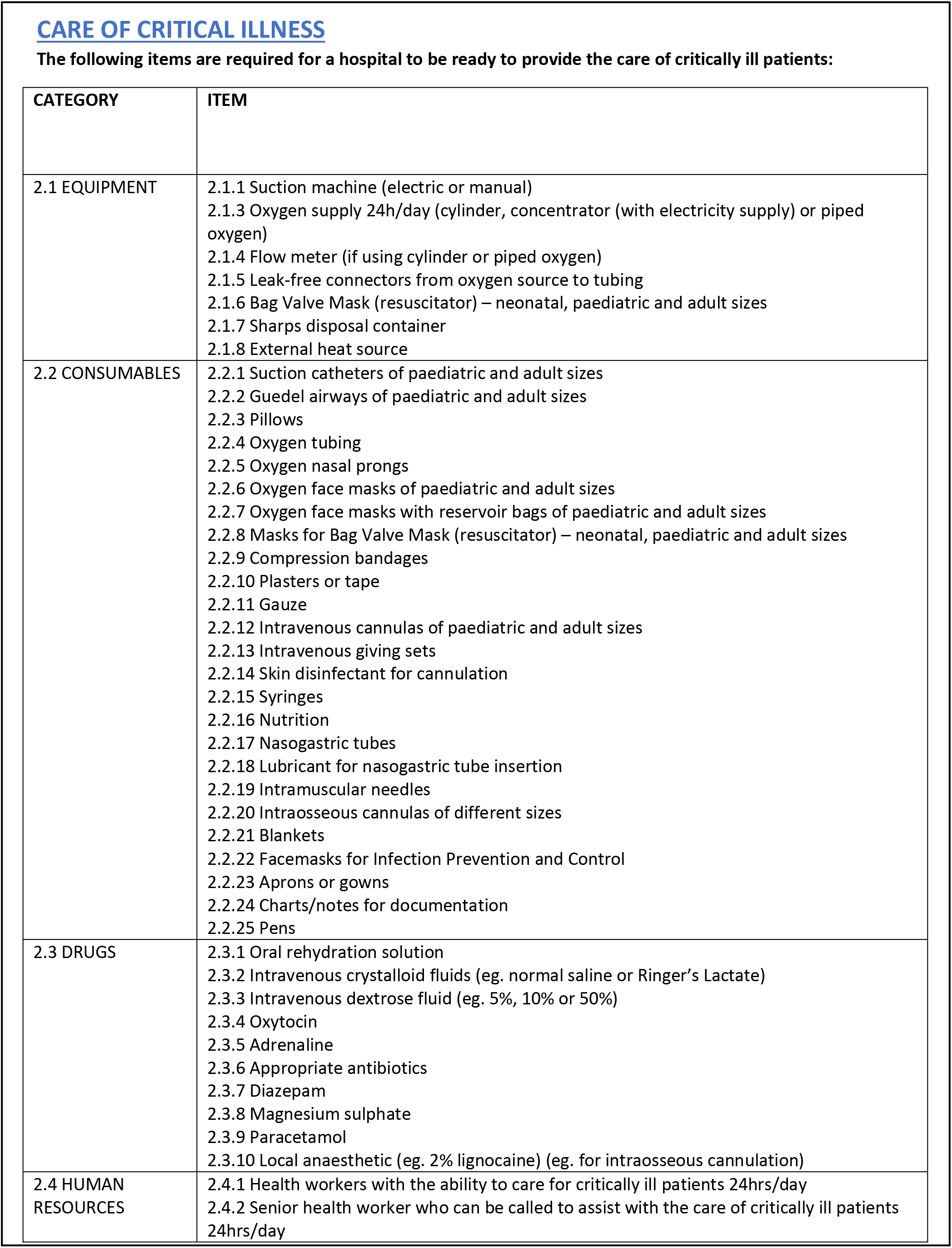
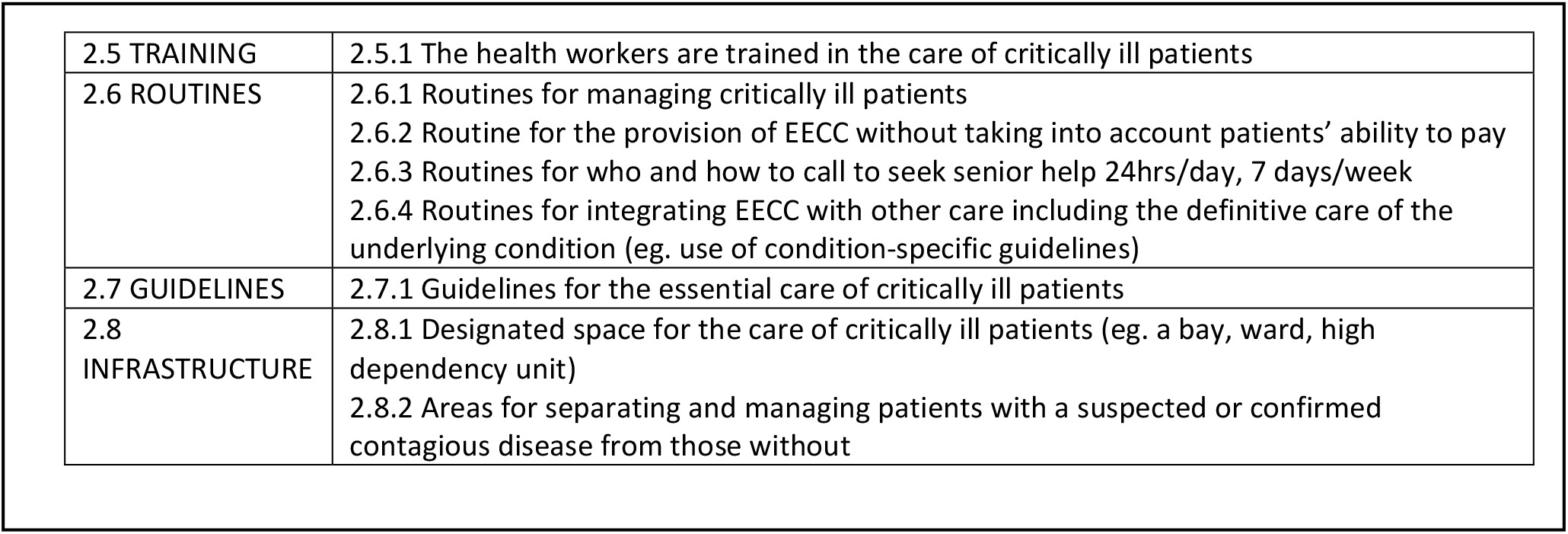
The Hospital Readiness Requirements for Essential Emergency and Critical Care.

| CATEGORY | ITEM |
| --- | --- |
| 1.1. EQUIPMENT | 1.1.1 Clock with second hand<br>1.1.2 Pulse oximeter & probe<br>1.1.3 Blood pressure measuring equipment (eg. sphygmomanometer with a stethoscope)<br>1.1.4 Blood pressure cuffs of different paediatric and adult sizes<br>1.1.5 Light source (lamp or flashlight)<br>1.1.6 Thermometer |
| 1.2 CONSUMABLES | 1.2.1 Soap or hand disinfectant<br>1.2.2 Examination gloves |
| 1.3 DRUGS | None |
| 1.4 HUMAN RESOURCES | 1.4.1 Health workers with the ability to identify critical illness 24h/day |
| 1.5 TRAINING | 1.5.1 The health workers are trained in the identification of critical illness |
| 1.6 ROUTINES | 1.6.1 Routines for the identification of critical illness |
| 1.7 GUIDELINES | 1.7.1 Guidelines for the identification of critical illness |
| 1.8 INFRASTRUCTURE | 1.8.1 Designated triage area (area for the identification of critical illness) in the Out-Patient Department or Emergency Unit (area of the hospital where patients arrive)<br>1.8.2 Running water |
| 2.1 EQUIPMENT | 2.1.1 Suction machine (electric or manual)<br>2.1.3 Oxygen supply 24h/day (cylinder, concentrator (with electricity supply) or piped oxygen)<br>2.1.4 Flow meter (if using cylinder or piped oxygen)<br>2.1.5 Leak-free connectors from oxygen source to tubing<br>2.1.6 Bag Valve Mask (resuscitator) – neonatal, paediatric and adult sizes<br>2.1.7 Sharps disposal container<br>2.1.8 External heat source |
| 2.2 CONSUMABLES | 2.2.1 Suction catheters of paediatric and adult sizes<br>2.2.2 Guedel airways of paediatric and adult sizes<br>2.2.3 Pillows<br>2.2.4 Oxygen tubing<br>2.2.5 Oxygen nasal prongs<br>2.2.6 Oxygen face masks of paediatric and adult sizes<br>2.2.7 Oxygen face masks with reservoir bags of paediatric and adult sizes<br>2.2.8 Masks for Bag Valve Mask (resuscitator) – neonatal, paediatric and adult sizes<br>2.2.9 Compression bandages<br>2.2.10 Plasters or tape<br>2.2.11 Gauze<br>2.2.12 Intravenous cannulas of paediatric and adult sizes<br>2.2.13 Intravenous giving sets<br>2.2.14 Skin disinfectant for cannulation<br>2.2.15 Syringes<br>2.2.16 Nutrition<br>2.2.17 Nasogastric tubes<br>2.2.18 Lubricant for nasogastric tube insertion<br>2.2.19 Intramuscular needles<br>2.2.20 Intraosseous cannulas of different sizes<br>2.2.21 Blankets<br>2.2.22 Facemasks for Infection Prevention and Control<br>2.2.23 Aprons or gowns<br>2.2.24 Charts/notes for documentation<br>2.2.25 Pens |
| 2.3 DRUGS | 2.3.1 Oral rehydration solution<br>2.3.2 Intravenous crystalloid fluids (eg. normal saline or Ringer's Lactate)<br>2.3.3 Intravenous dextrose fluid (eg. 5%, 10% or 50%)<br>2.3.4 Oxytocin<br>2.3.5 Adrenaline<br>2.3.6 Appropriate antibiotics<br>2.3.7 Diazepam<br>2.3.8 Magnesium sulphate<br>2.3.9 Paracetamol<br>2.3.10 Local anaesthetic (eg. 2% lignocaine) (eg. for intraosseous cannulation) |
| 2.4 HUMAN RESOURCES | 2.4.1 Health workers with the ability to care for critically ill patients 24hrs/day<br>2.4.2 Senior health worker who can be called to assist with the care of critically ill patients 24hrs/day |
| 2.5 TRAINING | 2.5.1 The health workers are trained in the care of critically ill patients |
| 2.6 ROUTINES | 2.6.1 Routines for managing critically ill patients<br>2.6.2 Routine for the provision of EECC without taking into account patients' ability to pay<br>2.6.3 Routines for who and how to call to seek senior help 24hrs/day, 7 days/week<br>2.6.4 Routines for integrating EECC with other care including the definitive care of the underlying condition (eg. use of condition-specific guidelines) |
| 2.7 GUIDELINES | 2.7.1 Guidelines for the essential care of critically ill patients |
| 2.8 INFRASTRUCTURE | 2.8.1 Designated space for the care of critically ill patients (eg. a bay, ward, high dependency unit)<br>2.8.2 Areas for separating and managing patients with a suspected or confirmed contagious disease from those without |

**PANEL 3.**
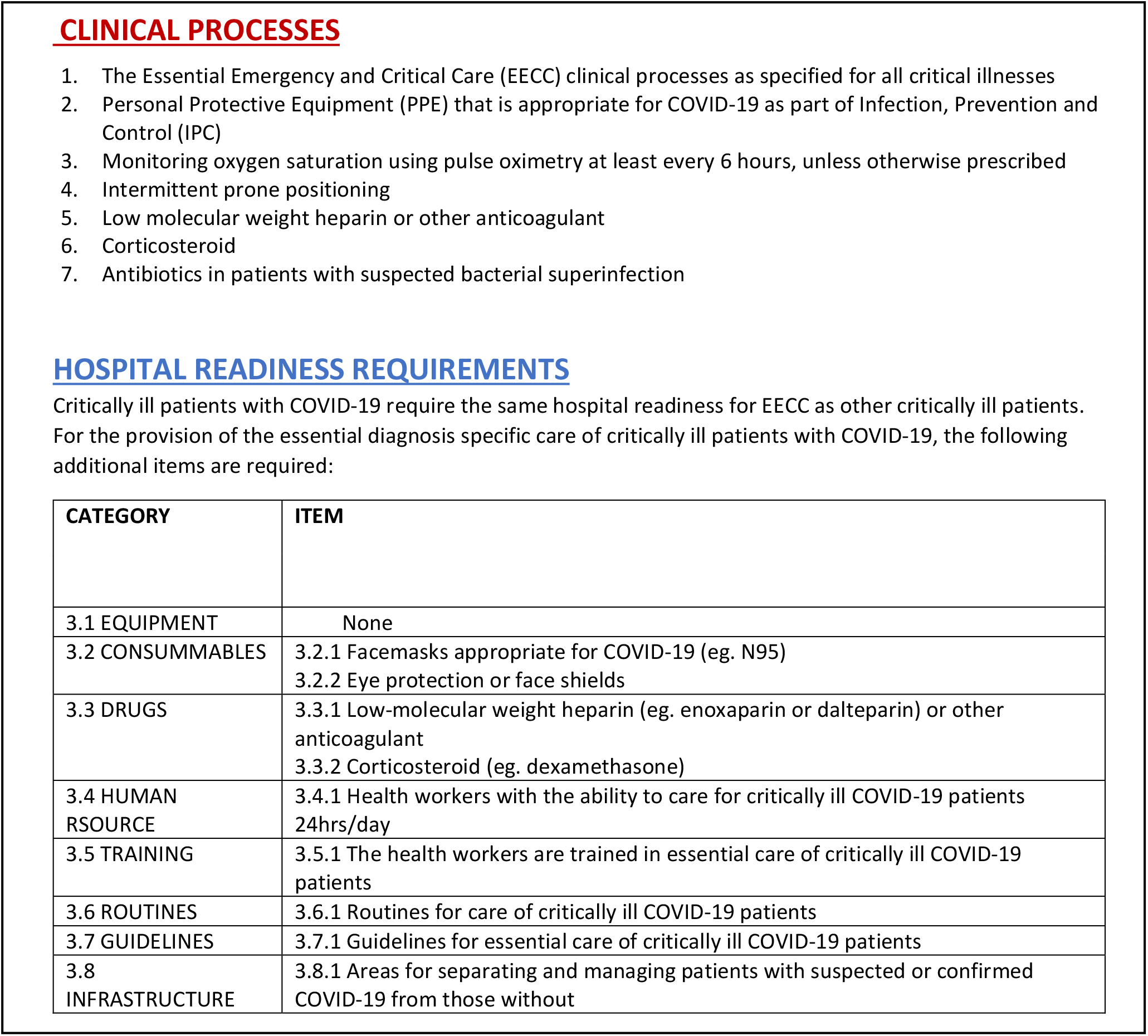
The essential diagnosis-specific care for critically ill patients with COVID-19.

| CATEGORY | ITEM |
| --- | --- |
| 3.1 EQUIPMENT | None |
| 3.2 CONSUMMABLES | 3.2.1 Facemasks appropriate for COVID-19 (eg. N95)<br>3.2.2 Eye protection or face shields |
| 3.3 DRUGS | 3.3.1 Low-molecular weight heparin (eg. enoxaparin or dalteparin) or other anticoagulant<br>3.3.2 Corticosteroid (eg. dexamethasone) |
| 3.4 HUMAN RESOURCE | 3.4.1 Health workers with the ability to care for critically ill COVID-19 patients 24hrs/day |
| 3.5 TRAINING | 3.5.1 The health workers are trained in essential care of critically ill COVID-19 patients |
| 3.6 ROUTINES | 3.6.1 Routines for care of critically ill COVID-19 patients |
| 3.7 GUIDELINES | 3.7.1 Guidelines for essential care of critically ill COVID-19 patients |
| 3.8 INFRASTRUCTURE | 3.8.1 Areas for separating and managing patients with suspected or confirmed COVID-19 from those without |

## Discussion

We have specified the content of essential emergency and critical care (EECC) based on consensus among global clinical experts. While the EECC approach is new, the included clinical processes are commonly used in the care of sick patients and can be seen in WHO publications and specialist society standards and guidelines [29, 32-35, 38, 39, 45]. The contribution of this study is the specification of a baseline bundle of care interventions that should be provided when needed to all critically ill patients in all hospitals in the world. This marks a break from previous guidelines that tend to be specialty-specific, condition-specific or location-specific, or that specify care that may be too complex and costly to provide in all hospital settings.

### The EECC approach

EECC is an approach that supports priority-setting in health systems. In this regard, it has parallels to the approaches used in the WHO’s Essential Medicines List [37], Interagency Integrated Triage Tool [29], Emergency Triage and Treatment for Children [32] and Universal Health Coverage [36]. EECC emphasises the identification and care of the critically ill, and the provision of the life-saving supportive care that is of low cost and of low complexity [25]. EECC can be seen as a unifying concept for such aspects of patient management found in WHO and specialist guidelines, triage, early warning systems and rapid response teams [28, 29, 39, 46, 47]. To maintain focus on life-saving supportive care and to be useful across all specialties, EECC does not include the definitive care of the underlying diagnoses. Instead, EECC is intended to complement speciality-based care and existing guidelines and does not aim to include all the care a patient needs – as well as EECC, patients should receive diagnostics, definitive and symptomatic care of their condition, additional nursing care, and if available, higher levels of emergency and critical care. It seeks to bridge the quality gap that is commonly found between the current care of critical illness and best-practice guidelines [12, 48, 49]. To ensure feasibility in settings with restricted human resources, EECC is designed to enable task-sharing between health professionals [50]. It should be noted that not all the EECC clinical processes will be needed in the care of every critically ill patient – they should be seen as essential “tools in the tool-box**”** for health workers to use when required. To operationalise the EECC approach, it is intended that the content specified here is used to develop tools for quality monitoring, teaching and integration into other guidelines and recommendations.

### EECC complements the current healthcare organisation

The basic clinical processes specified in EECC have been overlooked in healthcare [11, 13-15, 18, 20, 51, 52] In UK hospitals, half of patients received substandard basic vital organ support prior to intensive care and 31% of preventable deaths were associated with absent clinical monitoring [13, 14]. In Malawi, 89% of adult hypoxic patients, and 75% of children dying from pneumonia in hospital did not receive oxygen [17, 18]. The usual organisational set-up of health services may be one underlying reason for this. Specialist units with a primary function of delivering the definitive management for one disease group may under-estimate the effort needed to maintain core processes and competences in the supportive management of critically ill patients. Innovative and specialised treatments and technologies may become preferred to those that are basic and long-standing [53]. By targeting a feasible, lowest baseline quality for critically ill patients throughout hospital settings, EECC provides a complimentary approach to the current organisation that safeguards the provision of basic life-saving actions, enhancing the impact of hospital care for all acute conditions.

### EECC in the COVID-19 pandemic

EECC has added importance in a situation causing a substantial amount of severe disease and the Delphi panel agreed that EECC should be part of the care of critically ill patients with COVID-19. In addition, the agreed essential diagnostic-specific care for COVID-19 can assist in decisions about the priorities of care when the pandemic threatens to overwhelm available resources. All of the COVID-19 specific processes are well established and are included in the WHO COVID-19 clinical management guidance [30]. The WHO guidance, and others [54], additionally include recommendations for advanced critical care (such as mechanical ventilation, vasopressors and extracorporeal oxygenation), which may be difficult to rapidly scale-up in settings of low resources. Advanced critical care can be necessary to save the lives of some patients, but has a high cost per recovery and risks diverting scarce resources to a few individuals [55-60]. Fortunately the focus has shifted in the global pandemic response from advanced critical care towards securing basic oxygen delivery systems [61, 62] underscored by statements from the WHO and other partners [63, 64]. The impact of this shift, in and beyond the pandemic, could be even greater if the necessary processes for the effective use of oxygen and other care specified in EECC were included in the scale-up.

### Strengths and limitations

Our use of a consensus method with a large expert panel from diverse clinical and resource settings, specialties, and geographical locations gives the specified content legitimacy. The high response rate for this type of study during an ongoing pandemic illustrates the interest that experts had in the project’s aims. The high level of consensus (>90%) for the included clinical processes promotes confidence in the final package. However, the Delphi method does have limitations. It is expert-opinion based and is limited by the make-up of the panel. Only English language speakers were included, experts were not included from all countries, and the expedited timeline of the project due to the need for results that could impact the global response to the COVID-19 pandemic may have excluded experts who could have provided additional input. The initial content presented to the panel was aligned with WHO initiatives [39], and developed by a diverse specialist team, but the possibility remains that alternative methods would have led to a different output. The study did not address the underlying evidence-base for the included clinical processes, the impact, or the potential opportunity costs of increasing the coverage of EECC in hospitals – such system-wide effects warrant careful evaluation during EECC implementation. It should be noted that, while policy makers were involved throughout the process, the EECC content has not been ratified by the WHO or governmental ministries of health – the method has been primarily scientific. The findings should be seen as the first version of the EECC content, as recommended by global clinicians and researchers, one that could be incorporated into WHO and other global and national programmes and that should subsequently be improved and updated as new knowledge arises.

### Implications

Implementation of EECC could be an effective strategy as part of the current calls to save lives through improved quality of care in health systems [65] – a “low-hanging fruit”. Critically ill patients have high mortality rates in all hospital settings, especially where trained staff or resources are limited, and even small improvements in outcomes would have a large impact. EECC has a vital role in the ongoing COVID-19 pandemic, for the care of the surge of critically ill patients and for optimising the impact of the efforts to scale-up oxygen. Policy makers at global, national and regional levels aiming to reduce preventable deaths should focus on improved coverage of EECC and inclusion of EECC as part of universal health coverage[36].

## Conclusion

The content of essential emergency and critical care – and the essential care of critically ill patients with COVID-19 – has been specified using an inclusive global consensus. The content consists of effective, low-cost, and low-complexity life-saving care that is still frequently overlooked. The time has come to ensure that all patients in the world receive this care.

## Data Availability

Data available in supplementary tables

## Declarations

### Ethics approval and consent to participate

The study was approved by the London School of Hygiene and Tropical Medicine Research Ethics Committee (Ref: 22575). All participants in the Delphi panel provided written consent.

### Author contributions

COS conceptualised and designed the study, acquired and analysed the data, and developed the first draft of the manuscript. TB and KK conceptualised and designed the study, and acquired and analysed the data. AWS and MJ contributed to the design, acquisition and analysis of data. LAW provided input into study design and JNO, HRS, NR, JCM, JR, CH and RKK contributed to the design and analysis. All the authors interpreted the findings, critically revised the manuscript, and approved the final version.

## Acknowledgements

We thank the World Federation of Intensive and Critical Care (WFICC) for supporting the study. We also thank all experts who participated in the Delphi panel, those who took part in piloting the questions, and Anders Wennerstrand for communications advice.

## Abbreviations

ACVPU: Alert, confusion, voice, pain, unresponsive AVPU Alert, voice, pain, unresponsive
EECC: Essential Emergency and Critical Care
ICRC: The International Committee of the Red Cross
IPC: Infection prevention and control
SpO2: Saturation of haemoglobin with oxygen as measured by pulse oximetry
T&A: Treatments and Actions
WHO: The World Health Organization

## Footnotes

* *Effectiveness:* Established or proven to be safe and to reduce mortality. (compression to stop bleeding is effective; treating with leech therapy is not) *Feasibility:* Low-cost and low complexity. Possible to provide in a low-staffed, low-resourced setting without the immediate presence of a doctor (placing a comatose patient in the recovery position (lateral position) is feasible; continuous haemodialysis is not). *Universality:* Supports vital organ function rather than being the definitive care of a diagnosis. (IV fluids for shock are universal; thrombolytic therapy is not). *Relevance:* Established or proven to be a treatment for COVID-19.

