## Supplementary Tables for "Essential Emergency and Critical Care – a consensus among global clinical experts"

### Supplementary material

#### Supplementary Table 1. Delphi Results

*Level of agreement among Delphi experts in each round for the treatments and actions. Agreement is defined as the proportion of experts that rated "strongly agree" or "agree" out of those who provided a rating. Consensus is defined as ≥90% agreement (green), and non-consensus is <90% (orange).*
*EECC; Essential Emergency and Critical care. AVPU; Alert, Voice, Pain, Unresponsive.*

| Treatments and Actions | Round 1 N=269 Agreement (%) | Round 2 N=228 Agreement (%) | Round 3 N=194 Agreement (%) |
|---|---|---|---|
| 1. Identification of critical illness in EECC includes evaluating a limited number of physiological signs. The physiological signs used in EECC should include: | | | |
| The overall condition of the patient (concern that the patient is critically ill) | 96.6 | | |
| Presence of abnormal airway sounds (eg. snoring, gurgling, stridor) | 97.0 | | |
| Respiratory rate | 99.3 | | |
| Oxygen saturation (SpO2) | 95.9 | | |
| Pulse rate | 99.6 | | |
| Blood pressure | 95.9 | | |
| Level of consciousness (eg. "AVPU" or Glasgow Coma Scale) | 99.3 | | |
| Temperature | 88.0 | 85.9 | 93.3 |
| Capillary refill time | | 84.0 | 94.8 |
| Cold or warm extremities | | 80.9 | 83.0 |
| Cyanosis | | 82.3 | 87.5 |
| Confused, agitated or disoriented mental state | | 89.9 | 94.3 |
| Presence of prostration or lethargy | | 77.5 | 83.4 |
| Inability to stand or walk without help | | 63.1 | 64.8 |
| Presence of a generalized seizure | | 82.1 | 91.1 |

| | | | |
|---|---|---|---|
| Presence of respiratory distress (eg unable to complete sentences; accessory muscle use; chest recessions; grunting or head nodding) | | 93.4 | |
| Presence of severe dehydration (eg decreased skin turgor; dry mucous membranes; sunken fontanelle) | | 91.2 | |
| Inability to breastfeed or feed in a young child | | 86.0 | 92.9 |
| Presence of severe acute malnutrition | | 72.8 | 83.8 |
| 2. An evaluation of physiological signs should be conducted at these times: | | | |
| When a patient arrives at hospital seeking acute care | 100 | | |
| When a health worker is concerned that a patient may be critically ill | 98.9 | | |
| For hospital in-patients, at least every 24 hours, unless otherwise prescribed (Note: can be done more frequently) | 86.9 | 89.4 | 96.4 |
| More frequently for patients who are at risk of becoming critically ill or who are critically ill (eg. every 12hrs, every 6 hrs ... etc) | 99.6 | | |
| Less frequently for patients who have improved and are now stable (eg ...every 6hrs, every 12hrs, every 24hrs) | 84.4 | 95.6 | |
| Following a treatment or action (re-evaluation) | 98.9 | | |
| When a patient, family member or guardian is concerned that the patient may be critically ill | | 88.6 | 94.8 |
| During and after transport/transfer of a patient who is critically ill or at risk of becoming critically ill | | 98.3 | |
| During and after surgery or anaesthesia | | 95.6 | |

| Treatments and Actions | Round 1 N=269 Agreement (%) | Round 2 N=228 Agreement (%) | Round 3 N=194 Agreement (%) |
|---|---|---|---|
| 3. Communication about the patient's physiological signs in EECC should include: | | | |
| Documentation of the vital signs in the patient notes | 99.6 | | |
| Documentation in the patient notes when critical illness has been identified | 98.5 | | |
| Clear communication within the care team that a patient is critically ill (eg. verbal communication, at staff handovers, visible colour-coding) | 99.6 | | |
| 4. EECC includes treatment & actions when a patient has a blocked or threatened airway. The treatments & actions for a blocked or threatened airway should include: | | | |
| Recovery position (lateral position) | 91.3 | | |
| Age-appropriate airway positioning (eg chin lift or jaw thrust in adults, neutral position in young children) | 99.6 | | |
| Removal of any visible foreign body from the mouth | 98.1 | | |
| Suction for secretions that are obstructing the airway | 97.8 | | |
| Oro-pharyngeal (Guedel) airway | 92.1 | | |
| Naso-pharyngeal airway | 85.4 | 84.4 | 83.7 |
| Age-appropriate chest thrusts/ abdominal thrusts/ back blows in choking | 94.3 | | |

| 5. EECC includes treatments & actions when a patient has respiratory distress or hypoxia. The treatments & actions for respiratory distress or hypoxia should include: | | | |
|---|---|---|---|
| Optimize patient position (eg. sitting-up or prone) | 99.3 | | |
| Oxygen therapy using nasal prongs | 90.0 | 91.7 | |
| Oxygen therapy using facemask | 97.4 | | |
| Oxygen therapy using a mask with a reservoir bag (non re-breathing mask) | 95.8 | | |
| Breathing exercises (eg. deep breaths, coughing, changing position, expiration against mild resistance) | 78.5 | 76.4 | 72.1 |
| Inhaled bronchodilators (eg salbutamol) | 82.8 | 84.0 | 86.5 |
| Bag-valve-mask ventilation for newborns | 97.9 | | |
| Bag-valve-mask ventilation for children | | 91.3 | |
| Bag-valve-mask ventilation for adults | | 86.7 | 96.8 |

| Treatments and Actions | Round 1 N=269 Agreement (%) | Round 2 N=228 Agreement (%) | Round 3 N=194 Agreement (%) |
|---|---|---|---|
| 6. EECC includes treatments & actions when a patient has a threatened circulation or shock. The treatments & actions for a threatened circulation or shock should include: | | | |
| Optimise patient position (eg. lying flat, head-down, raised-legs, lateral tilt in pregnancy) | 99.3 | | |
| Compression and elevation to stop bleeding | 99.6 | | |
| Uterine massage when indicated | 97.9 | | |
| Appropriate bolus of intravenous fluid | 98.5 | | |
| Oral rehydration solution or other appropriate oral fluids for dehydration without shock | 81.0 | 94.3 | |
| Intramuscular adrenaline for anaphylaxis | | 90.6 | |
| Blood transfusion | | 75.5 | 85.3 |
| Tourniquet in severe limb bleeding† | | 74.2 | 88.2 |
| Appropriate antibiotics for sepsis | | 97.4 | |
| Oxytocin when indicated | | 96.2 | |
| Tranexamic Acid for haemorrhage | | 79.1 | 83.1 |
| 7. EECC includes treatments & actions when a patient has a reduced level of consciousness. The treatments & actions for a reduced level of consciousness should include: | | | |
| Treating an unconscious patient as having a threatened airway | 99.3 | | |
| Dextrose (iv or buccal) in unconsciousness or seizures unless bedside blood glucose testing rules out hypoglycaemia or there is a clear alternative cause | 92.8 | | |
| Protecting patients with a seizure from harm | 98.1 | | |

| | |
|---|---|
| Quick-acting anti-seizure medication for prolonged seizures (eg. iv or rectal diazepam) | 98.9 |
| Quick-acting anti-seizure medication for seizures in pregnancy and post-partum (eg. im magnesium sulphate) | 99.2 |
| Cooling (eg. fans, antipyretics) in severe hyperthermia with a reduced level of consciousness. | 93.5 |

| Treatments and Actions | Round 1 N=269 Agreement (%) | Round 2 N=228 Agreement (%) | Round 3 N=194 Agreement (%) |
|---|---|---|---|
| 8. EECC includes additional clinical treatments & actions for critical illness. These clinical treatments & actions should be included in EECC: | | | |
| Use of other indicated EECC treatments & actions if there is poor response to treatment, or if the patient deteriorates | 98.8 | | |
| Discontinuation of a treatment or action that is no longer indicated if a patient improves | 94.7 | | |
| Discontinuation of a treatment or action that has been deemed to no longer be in the patient's best interest | 94.7 | | |
| Use of blankets and other means (including skin-to-skin care for babies) to keep the patient warm | 99.2 | | |
| Feeding (including breastfeeding for babies). naso-gastric feeding and dextrose for nutrition and to avoid hypoglycaemia | 98.1 | | |
| Treatment of pain and anxiety (eg. with needs-based psychological support, medication) | 98.9 | | |
| Mobilise the patient as early as possible | | 91.1 | |
| Insertion of an intravenous cannula when critical illness is identified | | 98.2 | |
| Insertion of an intraosseous cannula, if indicated, if an IV cannula is not possible | | 90.9 | |
| Regular turning of immobilised patients | | 94.3 | |
| Cervical spine stabilization in possible cervical spine injury | | 96.9 | |

| 9. EECC includes additional treatments & actions for critical illness. These additional treatments & actions should be included in EECC: | | | |
|---|---|---|---|
| Infection, prevention and control (IPC) measures including hand hygiene and separation of patients with a suspected or confirmed contagious disease from those without | 98.9 | | |
| Seeking of assistance from additional or senior staff when a critically ill patient is identified | 99.3 | | |
| Care for all critically ill patients in locations that facilitate observation and care (eg. designated beds, a bay or a unit for critically ill patients) | 99.6 | | |
| Respectful and patient-centred care | 99.3 | | |
| Effective and respectful communication with the patient and family | 99.3 | | |
| Provision of EECC without considering the patient's ability to pay | 97.0 | | |
| Clear documentation of all treatments & actions | 99.6 | | |
| Recognition when EECC is not sufficient to manage the critical illness | 98.9 | | |
| Integration with care that is outside the scope of EECC (eg. the need for prompt investigations, definitive treatment of underlying conditions, end-of-life care, referral) | 98.9 | | |
| Clear communication and documentation about the planned EECC (eg continue oxygen therapy, give IV fluids | | 97.4 | |
| Prevent delirium (eg. sleep hygiene, provision of the patient's glasses or hearing aid) | | 84.9 | 91.6 |

| Treatments and Actions | Round 1 N=269 Agreement (%) | Round 2 N=228 Agreement (%) | Round 3 N=194 Agreement (%) |
|---|---|---|---|
| 10. The essential care of a critically ill COVID-19 patient should include: | | | |
| The EECC treatments & actions as specified for all critical illnesses | 98.9 | | |
| Personal Protective Equipment (PPE) that is appropriate for COVID-19 as part of Infection, Prevention and Control (IPC) | 99.6 | | |
| Monitoring oxygen saturation using pulse oximetry at least every 6 hours, unless otherwise prescribed | 91.7 | | |
| Intermittent prone positioning | 90.9 | | |
| Low molecular weight heparin or other anticoagulant | 92.2 | | |
| Corticosteroid | 93.8 | | |
| Antibiotics in patients with suspected bacterial superinfection | 98.5 | | |
| Zinc | | 51.5 | 42.8 |
| Aspirin | | 54.5 | 57.1 |

**Supplementary Table 2 Subgroup analysis Round One**

*Agreement in the sub-groups of experts for the treatments and actions that reached consensus in Round One. Agreement is defined as the proportion of experts that rated "strongly agree" or "agree" out of those who provided a rating. Consensus is ≥90% agreement.*

| Reached Consensus in Round 1 | Delphi Panel | Intensive Care | | Emergency Care | | Low Income Country | | Doctor | |
|---|---|---|---|---|---|---|---|---|---|
| **Treatments and Actions** | | Intensive Care | Non-Intensive Care | Emergency Care | Non-Emergency Care | LICs | Non-LICs | Doctor | Non-doctor |
| 1.1 The overall condition of the patient (concern that the patient is critically ill) | 96.6 | 96.9 | 96.2 | 97.9 | 96.0 | 95.5 | 98.9 | 96.7 | 96.5 |
| 1.2 Presence of abnormal airway sounds (eg. snoring, gurgling, stridor) | 97.0 | 96.3 | 98.7 | 98.9 | 96.0 | 98.9 | 93.5 | 96.7 | 98.3 |
| 1.3 Respiratory rate | 99.3 | 99.5 | 98.7 | 98.9 | 99.4 | 99.4 | 98.9 | 99.1 | 100 |
| 1.4 Oxygen saturation (SpO2) | 95.9 | 96.3 | 94.8 | 93.6 | 97.1 | 97.1 | 93.5 | 94.8 | 100 |
| 1.5 Pulse rate | 99.6 | 100 | 98.7 | 98.9 | 100 | 99.4 | 100 | 99.5 | 100 |
| 1.6 Blood pressure | 95.9 | 96.3 | 94.8 | 95.7 | 96.0 | 96.0 | 95.7 | 94.8 | 100 |
| 1.7 Level of consciousness (eg. "AVPU" or Glasgow Coma Scale) | 99.3 | 99.5 | 98.7 | 98.9 | 99.4 | 98.9 | 100 | 99.1 | 100 |
| 2.1 When a patient arrives at hospital seeking acute care | 100 | 100 | 100 | 100 | 100 | 100 | 100 | 100 | 100 |
| 2.2 When a health worker is concerned that a patient may be critically ill | 98.9 | 99.0 | 98.7 | 98.9 | 98.9 | 98.9 | 98.9 | 99.1 | 98.3 |
| 2.6 Following a treatment or action (re-evaluation) | 98.9 | 98.9 | 98.7 | 98.9 | 98.9 | 98.3 | 100 | 99.1 | 98.3 |
| 3.1 Documentation of the vital signs in the patient notes | 99.6 | 99.5 | 100 | 100 | 99.4 | 99.4 | 100 | 99.5 | 100 |
| 3.2 Documentation in the patient notes when critical illness has been identified | 98.5 | 98.4 | 98.7 | 97.9 | 98.9 | 97.7 | 100 | 99.1 | 96.5 |
| 3.3 Clear communication within the care team that a patient is critically ill (eg. verbal communication, at staff handovers, visible colour-coding) | 99.6 | 99.5 | 100 | 98.9 | 100 | 99.4 | 100 | 100 | 98.3 |
| 4.1 Recovery position (lateral position) | 91.3 | 91.4 | 91.0 | 91.1 | 91.4 | 90.4 | 93.2 | 92.4 | 87.3 |
| 4.2 Age-appropriate airway positioning (eg chin lift or jaw thrust in adults, neutral position in young children) | 99.6 | 99.5 | 100 | 100 | 99.4 | 100 | 98.9 | 99.5 | 100 |
| 4.3 Removal of any visible foreign body from the mouth | 98.1 | 97.3 | 100 | 97.9 | 98.3 | 97.7 | 98.9 | 97.6 | 100 |
| 4.4 Suction for secretions that are obstructing the airway | 97.8 | 98.4 | 96.2 | 97.9 | 97.7 | 97.2 | 98.9 | 97.2 | 100 |
| 4.5 Oro-pharyngeal (Guedel) airway | 92.1 | 92.1 | 92.1 | 87.0 | 94.8 | 93.7 | 89.0 | 90.9 | 96.4 |
| 4.7 Age-appropriate chest thrusts/ abdominal thrusts/ back blows in choking | 94.3 | 93.1 | 97.4 | 94.6 | 94.2 | 94.9 | 93.3 | 93.8 | 96.4 |
| 5.1 Optimize patient position (eg. sitting-up or prone) | 99.3 | 99.5 | 98.7 | 97.9 | 100 | 99.4 | 98.9 | 99.5 | 98.3 |

| | | | | | | | | | |
|---|---|---|---|---|---|---|---|---|---|
| 5.3 Oxygen therapy using facemask | 97.4 | 99.0 | 93.7 | 97.9 | 97.2 | 97.7 | 96.7 | 99.1 | 91.2 |
| 5.4 Oxygen therapy using a mask with a reservoir bag (non re-breathing mask) | 95.8 | 95.8 | 96.0 | 94.6 | 96.5 | 96.5 | 94.5 | 95.2 | 98.3 |
| 5.7 Bag-valve-mask ventilation for newborns | 97.9 | 97.6 | 98.6 | 97.7 | 98.0 | 98.2 | 97.4 | 97.8 | 98.2 |
| 6.1 Optimise patient position (eg. lying flat, head-down, raised-legs, lateral tilt in pregnancy) | 99.3 | 99.0 | 100 | 98.9 | 99.4 | 98.9 | 100 | 99.0 | 100 |
| 6.2 Compression and elevation to stop bleeding | 99.6 | 99.5 | 100 | 98.9 | 100 | 99.4 | 100 | 100 | 98.3 |
| 6.3 Uterine massage when indicated | 97.9 | 98.8 | 95.6 | 98.8 | 97.4 | 98.2 | 97.3 | 97.3 | 100 |
| 6.4 Appropriate bolus of intravenous fluid | 98.5 | 98.4 | 98.7 | 97.9 | 98.9 | 99.4 | 96.7 | 98.1 | 100 |
| 7.1 Treating an unconscious patient as having a threatened airway | 99.3 | 100 | 97.4 | 100 | 98.9 | 99.4 | 98.9 | 99.5 | 98.3 |
| 7.2 Dextrose (iv or buccal) in unconsciousness or seizures unless bedside blood glucose testing rules out hypoglycaemia or there is a clear alternative cause | 92.8 | 92.0 | 94.7 | 96.7 | 90.6 | 93.6 | 91.1 | 92.3 | 94.6 |
| 7.3 Protecting patients with a seizure from harm | 98.1 | 97.9 | 98.7 | 98.9 | 97.7 | 98.3 | 97.8 | 97.6 | 100 |
| 7.4 Quick-acting anti-seizure medication for prolonged seizures (eg. iv or rectal diazepam) | 98.9 | 98.4 | 100 | 97.8 | 99.4 | 99.4 | 97.8 | 98.6 | 100 |
| 7.5 Quick-acting anti-seizure medication for seizures in pregnancy and post-partum (eg. im magnesium sulphate) | 99.2 | 98.9 | 100 | 98.9 | 99.4 | 100 | 97.7 | 99.5 | 98.2 |
| 7.6 Cooling (eg. fans, antipyretics) in severe hyperthermia with a reduced level of consciousness. | 93.5 | 93.0 | 94.7 | 92.3 | 94.1 | 93.6 | 93.3 | 93.2 | 94.6 |
| 8.1 Use of other indicated EECC treatments & actions if there is poor response to treatment, or if the patient deteriorates | 98.8 | 98.9 | 98.6 | 98.9 | 98.8 | 98.2 | 100 | 98.5 | 100 |
| 8.2 Discontinuation of a treatment or action that is no longer indicated if a patient improves | 94.7 | 93.6 | 97.4 | 91.4 | 96.5 | 94.8 | 94.4 | 96.1 | 89.5 |
| 8.3 Discontinuation of a treatment or action that has been deemed to no longer be in the patient’s best interest | 94.7 | 94.7 | 94.7 | 92.4 | 96.0 | 94.3 | 95.6 | 96.2 | 89.5 |
| 8.4 Use of blankets and other means (including skin-to-skin care for babies) to keep the patient warm | 99.2 | 98.9 | 100 | 100 | 98.8 | 99.4 | 98.9 | 99.5 | 98.2 |
| 8.5 Feeding (including breastfeeding for babies), naso-gastric feeding and dextrose for nutrition and to avoid hypoglycaemia | 98.1 | 97.9 | 98.7 | 97.8 | 98.3 | 98.3 | 97.8 | 97.6 | 100 |
| 8.6 Treatment of pain and anxiety (eg. with needs-based psychological support, medication) | 98.9 | 99.0 | 98.7 | 98.9 | 98.9 | 98.3 | 100 | 98.6 | 100 |
| 9.1 Infection, prevention and control (IPC) measures including hand hygiene and separation of patients with a suspected or confirmed contagious disease from those without | 98.9 | 98.4 | 100 | 98.9 | 98.9 | 98.9 | 98.9 | 98.6 | 100 |
| 9.2 Seeking of assistance from additional or senior staff when a critically ill patient is identified | 99.3 | 99.0 | 100 | 98.9 | 99.4 | 100 | 97.8 | 99.1 | 100 |
| 9.3 Care for all critically ill patients in locations that facilitate observation and care (eg. designated beds, a bay or a unit for critically ill patients) | 99.6 | 99.5 | 100 | 100 | 99.4 | 99.4 | 100 | 99.5 | 100 |

| | | | | | | | | | |
|---|---|---|---|---|---|---|---|---|---|
| 9.4 Respectful and patient-centred care | 99.3 | 100 | 97.5 | 97.9 | 100 | 98.9 | 100 | 99.1 | 100 |
| 9.5 Effective and respectful communication with the patient and family | 99.3 | 100 | 97.5 | 97.9 | 100 | 98.9 | 100 | 99.1 | 100 |
| 9.6 Provision of EECC without considering the patient's ability to pay | 97.0 | 96.8 | 97.4 | 95.7 | 97.7 | 96.6 | 97.8 | 96.7 | 98.3 |
| 9.7 Clear documentation of all treatments & actions | 99.6 | 100 | 98.7 | 98.9 | 100 | 99.4 | 100 | 99.5 | 100 |
| 9.8 Recognition when EECC is not sufficient to manage the critical illness | 98.9 | 98.4 | 100 | 97.9 | 99.4 | 98.9 | 98.9 | 99.5 | 96.5 |
| 9.9 Integration with care that is outside the scope of EECC (eg. the need for prompt investigations, definitive treatment of underlying conditions, end-of-life care, referral) | 98.9 | 98.4 | 100 | 98.9 | 98.9 | 98.9 | 98.9 | 99.1 | 98.3 |
| 10.1 The EECC treatments & actions as specified for all critical illnesses | 98.9 | 98.4 | 100 | 97.8 | 99.4 | 99.4 | 97.8 | 99.0 | 98.2 |
| 10.2 Personal Protective Equipment (PPE) that is appropriate for COVID-19 as part of Infection, Prevention and Control (IPC) | 99.6 | 100 | 98.7 | 98.9 | 100 | 100 | 98.9 | 99.5 | 100 |
| 10.3 Monitoring oxygen saturation using pulse oximetry at least every 6 hours, unless otherwise prescribed | 91.7 | 90.5 | 94.7 | 87.0 | 94.2 | 90.2 | 94.5 | 92.4 | 89.1 |
| 10.4 Intermittent prone positioning | 90.9 | 88.5 | 97.1 | 94.1 | 89.3 | 94.6 | 84.1 | 89.4 | 96.4 |
| 10.5 Low molecular weight heparin or other anticoagulant | 92.2 | 91.2 | 95.2 | 92.8 | 91.9 | 91.9 | 92.9 | 90.6 | 98.1 |
| 10.6 Corticosteroid | 93.8 | 92.5 | 97.2 | 97.8 | 91.6 | 94.7 | 92.1 | 92.6 | 98.2 |
| 10.7 Antibiotics in patients with suspected bacterial superinfection | 98.5 | 98.4 | 98.7 | 97.8 | 98.9 | 97.7 | 100 | 99.0 | 96.5 |

**Supplementary Table 3 Subgroup analysis Round Two**

*Agreement (%) in subgroups for the treatments and actions that reached consensus in Round Two*

| Reached Consensus in Round 2 | Delphi Panel | Intensive Care | | Emergency Care | | Low Income Country | | Doctor | |
|---|---|---|---|---|---|---|---|---|---|
| **Treatments and Actions** | | Intensive Care | Non-Intensive Care | Emergency Care | Non-Emergency Care | LICs | Non-LICs | Doctor | Non-doctor |
| 1.16 Presence of respiratory distress (eg unable to complete sentences; accessory muscle use; chest recessions; grunting or head nodding) | 93.4 | 95.2 | 88.7 | 96.3 | 91.8 | 94.4 | 91.0 | 92.5 | 97.6 |
| 1.17 Presence of severe dehydration (eg decreased skin turgor; dry mucous membranes; sunken fontanelle) | 91.2 | 90.9 | 91.8 | 93.8 | 89.7 | 92.5 | 88.1 | 90.8 | 92.7 |
| 2.5 Less frequently for patients who have improved and are now stable (eg ...every 6hrs, every 12hrs, every 24hrs) | 95.6 | 95.2 | 96.8 | 95.0 | 96.0 | 95.7 | 95.5 | 96.8 | 90.2 |
| 2.8 During and after transport/transfer of a patient who is critically ill or at risk of becoming critically ill | 98.3 | 98.2 | 98.4 | 97.5 | 98.7 | 98.8 | 97.0 | 98.4 | 97.6 |
| 2.9 During and after surgery or anaesthesia | 95.6 | 95.8 | 95.1 | 96.3 | 95.2 | 97.5 | 90.8 | 95.1 | 97.6 |
| 5.2 Oxygen therapy using nasal prongs | 91.7 | 89.8 | 96.8 | 88.8 | 93.2 | 90.1 | 95.5 | 93.1 | 85.4 |
| 5.8 Bag-valve-mask ventilation for children | 91.3 | 90.3 | 94.2 | 94.8 | 89.2 | 91.2 | 91.7 | 91.0 | 92.7 |
| 6.5 Oral rehydration solution or other appropriate oral fluids for dehydration without shock | 94.3 | 94.6 | 93.6 | 94.9 | 93.9 | 93.2 | 97.0 | 94.7 | 92.5 |
| 6.6 Intramuscular adrenaline for anaphylaxis | 90.6 | 91.3 | 88.7 | 92.3 | 89.7 | 88.6 | 95.4 | 92.0 | 83.3 |
| 6.9 Appropriate antibiotics for sepsis | 97.4 | 97.6 | 96.7 | 97.5 | 97.3 | 96.9 | 98.5 | 97.3 | 97.6 |
| 6.10 Oxytocin when indicated | 96.2 | 95.4 | 98.2 | 96.0 | 96.3 | 98.0 | 91.5 | 96.5 | 94.6 |
| 8.7 Mobilise the patient as early as possible | 91.1 | 92.1 | 88.3 | 95.0 | 88.9 | 93.1 | 86.2 | 89.1 | 100 |
| 8.8 Insertion of an intravenous cannula when critical illness is identified | 98.2 | 98.8 | 96.8 | 98.7 | 98.0 | 97.5 | 100 | 97.9 | 100 |
| 8.9 Insertion of an intraosseous cannula, if indicated, if an IV cannula is not possible | 90.9 | 92.6 | 86.2 | 93.6 | 89.4 | 91.1 | 90.3 | 90.6 | 92.3 |
| 8.10 Regular turning of immobilised patients | 94.3 | 94.0 | 95.2 | 93.8 | 94.6 | 94.4 | 94.0 | 93.1 | 100 |
| 8.11 Cervical spine stabilization in possible cervical spine injury | 96.9 | 96.4 | 98.4 | 97.5 | 96.6 | 97.5 | 95.5 | 96.8 | 97.6 |
| 9.10 Clear communication and documentation about the planned EECC (eg continue oxygen therapy, give IV fluids | 97.4 | 96.4 | 100 | 98.8 | 96.6 | 98.1 | 95.5 | 96.8 | 100 |

**Supplementary Table 4: Subgroup analysis Round Three**

*Agreement (%) in subgroups for the treatments and actions that reached consensus in Round 3*

| Reached Consensus in Round 3 | Delphi Panel | Intensive Care | | Emergency Care | | Low Income Country | | Doctor | |
|---|---|---|---|---|---|---|---|---|---|
| **Treatments and Actions** | | Intensive Care | Non-Intensive Care | Emergency Care | Non-Emergency Care | LICs | Non-LICs | Doctor | Non-doctor |
| 1.8 Temperature | 93.3 | 92.8 | 94.6 | 94.0 | 92.9 | 93.5 | 92.9 | 93.1 | 94.3 |
| 1.9 Capillary refill time | 94.8 | 94.9 | 94.4 | 97.0 | 93.7 | 95.6 | 92.9 | 93.6 | 100 |
| 1.12 Confused, agitated or disoriented mental state | 94.3 | 94.2 | 94.6 | 97.0 | 92.9 | 94.9 | 92.9 | 93.1 | 100 |
| 1.15 Presence of a generalized seizure | 91.1 | 89.1 | 96.3 | 95.4 | 88.9 | 94.9 | 81.8 | 89.7 | 97.1 |
| 1.18 Inability to breastfeed or feed in a young child | 92.9 | 92.4 | 94.1 | 90.6 | 94.1 | 93.8 | 90.7 | 92.6 | 94.1 |
| 2.3 For hospital in-patients, at least every 24 hours, unless otherwise prescribed (Note: can be done more frequently) | 96.4 | 95.7 | 98.2 | 95.5 | 96.9 | 95.7 | 98.2 | 96.2 | 97.1 |
| 2.7 When a patient, family member or guardian is concerned that the patient may be critically ill | 94.8 | 94.9 | 94.6 | 94.0 | 95.2 | 95.7 | 92.7 | 95.6 | 91.4 |
| 5.9 Bag-valve-mask ventilation for adults | 96.8 | 96.4 | 98.1 | 98.5 | 95.9 | 96.3 | 98.2 | 96.1 | 100 |
| 9.11 Prevent delirium (eg. sleep hygiene, provision of the patient's glasses or hearing aid) | 91.6 | 92.1 | 90.2 | 90.8 | 92.0 | 91.2 | 92.6 | 91.0 | 94.3 |
